# Mapping evoked fields in primary motor and sensory areas via magnetoencephalography in tetraplegia

**DOI:** 10.1101/2021.05.01.21256017

**Authors:** Stephen Foldes, Santosh Chandrasekaran, Joseph Camerone, James Lowe, Richard Ramdeo, John Ebersole, Chad E Bouton

## Abstract

Devices interfacing with the brain through implantation in cortical or subcortical structures have great potential for restoration and rehabilitation in patients with sensory or motor dysfunction. Typical implantation surgeries are planned based on maps of brain activity generated from intact function. However, mapping brain activity for planning implantation surgeries is challenging in the target population due to abnormal residual function and, increasingly often, existing MRI-incompatible implanted hardware. Here, we present methods and results for mapping impaired somatosensory and motor function in an individual with paralysis and an existing brain-computer interface (BCI) device. Magnetoencephalography (MEG) was used to directly map the neural activity evoked during transcutaneous electrical stimulation and attempted movement of the impaired hand. Evoked fields were found to align with the expected anatomy and somatotopic organization. This approach may be valuable for guiding implants in other applications, such as cortical stimulation for pain and to improve implant targeting to help reduce the craniotomy size.

## 2 Introduction

In the United States over 5 million people are living with paralysis, with stroke and spinal cord injury (SCI) as the leading causes (Armour et al., 2016). Intracortical brain-computer interfaces (BCI) have been used to decode intended movements from neural activity recorded in the primary motor cortex (Ajiboye et al., 2017; Collinger et al., 2013; Gilja et al., 2015; Hochberg et al., 2012) and subsequently, used to restore intention-driven movement in tetraplegia (Bouton et al., 2016). To enable dexterous movements and decrease reliance on visual feedback, direct stimulation of the sensory cortex has recently been used to provide somatosensory feedback (Armenta Salas et al., 2018; Flesher et al., 2016). Cortical stimulation is also used for pain management (i.e. motor cortex stimulation - MCS) which may be valuable for people with paralysis who are known to have chronic pain (Mo et al., 2019). Cortical stimulation of somatosensory or motor areas may also be used in the future to enhance neural sprouting to improve therapeutic neuroplasticity (Carmel & Martin, 2014; Raineteau & Schwab, 2001; Xiong et al., 2017; Zareen et al., 2017). Despite the growth in the field of implanted neural interfaces, there is limited discussion determining the optimal planning and targeting of the implants.

Preparing for epilepsy or other respective brain surgeries in able-bodied patients typically entail pre-surgical mapping that relies on intact function, such as active movement. However, for people with impaired motor and sensory function standard mapping procedures are not always possible or optimal. Because this patient population is unable to move or perceive external somatosensory stimuli, mapping approaches for BCIs have typically been based on anatomy. Some studies have adapted standard pre-surgical mapping methods to help localize motor and sensory areas (Collinger et al., 2013; Collinger, Kryger, et al., 2014; Degenhart et al., 2018; Flesher et al., 2016).

For typical pre-surgical mapping, the goal is to determine eloquent areas to be avoided during surgery. The common methods are task-based fMRI, evoked field mapping with magnetoencephalography (MEG), or transcranial magnetic stimulation (TMS). Task-based fMRI identifies areas where blood flow changes when performing a task or processing a stimuli. This technique provides a 3D map of activation, but the signal has a multiple-seconds time scale leading to broad areas of activation (i.e. high sensitivity, but low specificity). Another caveat to fMRI is the necessity for strong magnetic RF fields which often incompatible with implants that a person with impairments often have (e.g. spinal bracing, peripheral nerve stimulation devices, etc.). TMS stimulates the brain transcutaneously to find cortical areas that initiate eloquent responses. Though TMS mapping is currently uncommon clinically, its most common application is for mapping motor function by sending magnetic pulse through the skull and activating the descending fibers of the motor cortex to induce a motor twitch at the hand. However, for many people with paralysis the connections from their brain to the hand are disrupted (i.e. spinal cord) impeding the TMS mapping. Furthermore, externally activating the brain with TMS induces activation of the white matter tracts related to function, though the target for most cortical BCIs is gray matter. TMS has been used in paralyzed populations but not pre-surgical planning (Defrin et al., 2007; Freund et al., 2011; Jo & Perez, 2020; Tazoe & Perez, 2015).

Contrary to fMRI and TMS, magnetoencephalography (MEG) directly maps the neural activity internally generated by the brain. MEG can also be performed safely for people who have metallic implants. Like all somatosensory and motor mapping, MEG mapping relies on the patient to have intact function. However, because cortical responses are expected to be weaker and have more complex patterns in people with known sensorimotor dysfunction (Foldes, Weber, & Collinger, 2017; Goto et al., 2002), traditional analysis methods are not ideal. For example, the standard dipole pole models used in traditional magnetic source imaging (i.e. equivalent current dipole – ECD) map single points to represent the brain activity during events, but are less accurate in cases of low signal-to-noise ratios and complex, widespread activation patterns (Hara et al., 2007; Shiraishi et al., 2005; Tanaka & Stufflebeam, 2014). Instead, distributed source models (DSM) overcome these limitations by modeling multiple dipoles across the brain surface to explain the recorded magnetic field. DSM map the amplitudes of the currents across the cortical surface that could have generated the recorded magnetic field.

Another limitation of traditional MEG mapping is the focus on early components of the evoked response. In particular, somatosensory evoked fields (SSEF) typically evaluate the activity 20 ms after electrical stimulation. This component of the response (i.e., N20m) originates from pyramidal cells in the posterior wall of the central sulcus (cortical area 3b). However, N20 response has a lower amplitude in patients with spinal cord injury compared to healthy controls likely due to afferent disruption (Goto et al., 2002). Later components of the evoked field (e.g., P100m) have stronger responses and map to areas that process more advanced information such as shapes and textures (Desmedt, Nguyen Tran Huy, & Bourguet, 1983). Furthermore, unlike the early components that correspond to the input signals, these later components correspond to the conscious experience which are the targets for BCIs (Dehaene & Naccache, 2001).

Optimized methods to map sensorimotor activity on the cortex are needed to guide brain interface devices of the future. These advanced mapping techniques are especially important for patients with sensorimotor impairment who have the most to gain from BCI technology. We present a method and results for mapping somatosensory and motor function in an individual with paralysis and an existing implanted BCI device. This approach may be valuable for guiding implants in other applications, such as cortical stimulation for the treatment of pain, and to improve implant targeting to help reduce the craniotomy size.

## 3 Methods

### 3.1 Participant

Sensorimotor mapping of left-hand function was performed for a participant in his late 20s with stable, non-spastic tetraplegia from cervical SCI sustained in a diving accident 8 years prior. The goal of the mapping was for surgical planning of a bidirectional BCI system. International Standards for Neurological Classification of Spinal Cord Injury (ISNCSCI) (Kirshblum et al., 2014) based assessment classified the participant’s neurologic level to C5 AIS A – motor complete (AIS: ASIA Impairment Scale; ASIA: American Spinal Injury Association) with zone of partial preservation mapped to C6 bilaterally. He had full bilateral shoulder and elbow flexion (grade 5/5) and active wrist extension with radial deviation, but with incomplete range of motion against gravity (grade 2/5). He had no motor function below the level of C6. While his neurologic sensory level was C6 on the left, an altered but present light touch on his thumb resulted in a level of C5 on the right. He had intact proprioception in the left upper limb at the shoulder for internal rotation through external rotation, at the elbow for flexion through extension, at the forearm for pronation through supination, and at the wrist for flexion through extension. Extension at the metacarpal-phalangeal joints was impaired for all digits.

The participant had been previously implanted with one Utah microelectrode array with 96, 1.5 mm long electrodes (Blackrock Microsystems) in the hand area of the left motor cortex (Bouton et al., 2016). The array was connected to a Blackrock NeuroPort pedestal made of titanium. MRI was not possible due to the existing implanted hardware. Procedures were performed under Northwell Health IRB Study #17-0840 and FDA-issued IDE #G170200, as detailed under Clinical Trials #NCT03680872. Data can be made available upon reasonable request.

### 3.2 MEG RECORDINGS

MEG data were recorded with a 306-channel whole-head system with 102 sensor-triplets containing a magnetometer, longitudinal gradiometer, and latitudinal gradiometer (Elekta Neuromag Vectorview). The participant was in a seated position. Head position in the MEG was recorded at the beginning of each run using localization coils. Raw MEG signals were band-pass filtered between 0.1 and 330 Hz and then sampled at 1000 Hz. MEG sensor data were preprocessed by manually removing bad channels prior to performing temporal signal-space separation (tSSS) with a 4 s buffer (Taulu & Hari, 2009). After visual inspection, 84 sensors over the left hemisphere were removed due to artifacts caused by metallic implants.

### 3.3 Tasks

Motor and somatosensory tasks were designed specifically for the participant’s intact function. The goal was to map left hand areas in the right sensorimotor cortex. To map the hand-motor functions that were completely impaired, the participant attempted and imagined flexing their left thumb or index finger in sync with the movement performed by an experimenter they were watching. The experimenter performing the movements was seated on the subject’s right side and performing movements with his left hand. Data were marked when the experimenter broke a light beam while performing thumb or index flexion (i.e. the data were marked by the experimenter, not the participant). The experimenter wore headphones to receive an audio cue to help maintain a steady rhythm of movement. An overt wrist extension was used to map the participants limited wrist motor function. For this task, an auditory cue indicated to the participant to extend their left wrist releasing a button to mark an event.

To map hand-sensory function, electrical stimulation was applied directly to proximal base of the left thumb (10.3 mA amplitude, 200 ms pulse duration) or between the middle and distal joint of the left index finger (9.1 mA amplitude, 200 ms pulse width) using a ring electrode (Natus Disposable Ring Adhesive Electrodes). A Digitimer DS7A delivered constant current stimulation. Due to residual sensation in the thumb, the participant could detect the thumb stimulation, but not the index finger stimulation. Two other evoked fields from intact function were mapped to confirm somatotopic organization. Somatosensory face area was also mapped from electrical stimulation (5.2 mA amplitude, 200 ms pulse duration) of the upper lip to evoke fully intact sensation. Inter-pulse interval for the electrical stimulation randomized between 2000-3000 ms and motor tasks were performed at an average inter-task interval of 1500 ms. The average frequency of the wrist movements was ranged from 1.6-2.8 Hz. Multiple runs of each task were performed consisting of >100 epochs each. Trial counts are shown in Table 1.

**Table 1:** Mapping parameters.

|  | Body Part | Task | Intact Function | # Trials | Component Information |  |  |
| --- | --- | --- | --- | --- | --- | --- | --- |
|  |  |  |  |  | Time (ms) | Peak amplitude (pA*m) | Rationale |
| Sensory | Thumb | eStim at D1 base | Limited residual sensation | 1431 | 42 | 3.7 | Early component at expected latency |
|  |  |  |  |  | 107 | 5.1 | Time series peak and during expected late-latency |
|  | Index | eStim at middle of D2 finger | No sensation | 808 | 30 | 7.7 | Peak in ROI at expected latency |
|  | Face | eStim at upper lip | Normal sensation | 808 | 26 | 25.1 | ROI time series peak at expected latency |
| Motor | Thumb | Attempted and imagined button pushing while observing actual pushing | No movement | 906 | 104 | 4.9 | Early peak in ROI, around reaction time |
|  | Index | Attempted and imagined button pushing while observing actual pushing | No movement | 722 | 67 | 9.4 | Early peak in ROI, around reaction time |
|  | Wrist | Actual extension to release button | Some residual motor function | 473 | -7 | 7.4 | Large peak before movement. Likely related to moving finger, not wrist |
|  |  |  |  |  | 58 | 8.3 | Clear peak post-trigger |

### 3.4 Source space transformation

MEG was transformed into source space using Brainstorm Toolbox (Tadel et al., 2011). For each experimental run, the head position within the MEG helmet was used to align the MEG sensor data with an anatomical model of the brain and head from the MRI (Gross et al., 2013). Forward models were generated using overlapping spheres method (Mosher, Leahy, & Lewis, 1999). Dipole sources were modeled on the brain surface and given three orientations (i.e. unconstrained cortical orientation). Removing the orthogonal source constraint allowed for gyral source activity to be modeled since the implantation target was known to be on the gyrus and not in the sulcus. This produced dipoles at 15,002 locations with 3 different orientations for a total of 45,006 dipoles. The inverse solution was computed using weighted minimum norm estimate (Tadel et al., 2011) with a noise covariance matrix calculated from pre-trigger data (−300 to -100 ms for motor tasks). The anatomical forward model was created from a whole-brain T1-weighted images with 1 mm3 voxels. Freesurfer was used to generate three dimensional models of the brain, CSF, skull, and scalp (Fischl, 2012).

### 3.5 Source Mapping

The average evoked fields generated from each task were evaluated independently. Because the data were noisy due to the participant’s implant/hardware and the evoked activity was diminished due to paralysis, special of considerations were taken to identify times used for mapping. The goal was to map somatosensory and motor areas on the gyrus to guide implantation of BCI electrodes. Therefore, the assessment of source activity was limited to regions of interest (ROIs) including the hand areas of the precentral gyrus for motor and postcentral gyrus for sensory. Evoked field waveforms were assessed for standard peaks looking in the right parietal MEG sensors as well as looking at the waveform of the maximal activity in the ROI. The source maps during the identified time period were evaluated for peak activity and for somatotopic organization. Intact function from wrist motor and face sensory areas were mapped to help confirm the somatotopic organization. The analysis of face sensory analysis used an ROI that included more medial post-central gyrus.

## 4 Results

Evoked activity was identified and mapped for all tasks, even though the motor and sensory capacity of the participant was impaired. Figure 3 shows the peak of the source maps and the broader organization. Table 1 shows the times and parameters used for source mapping.

**Figure 3:**
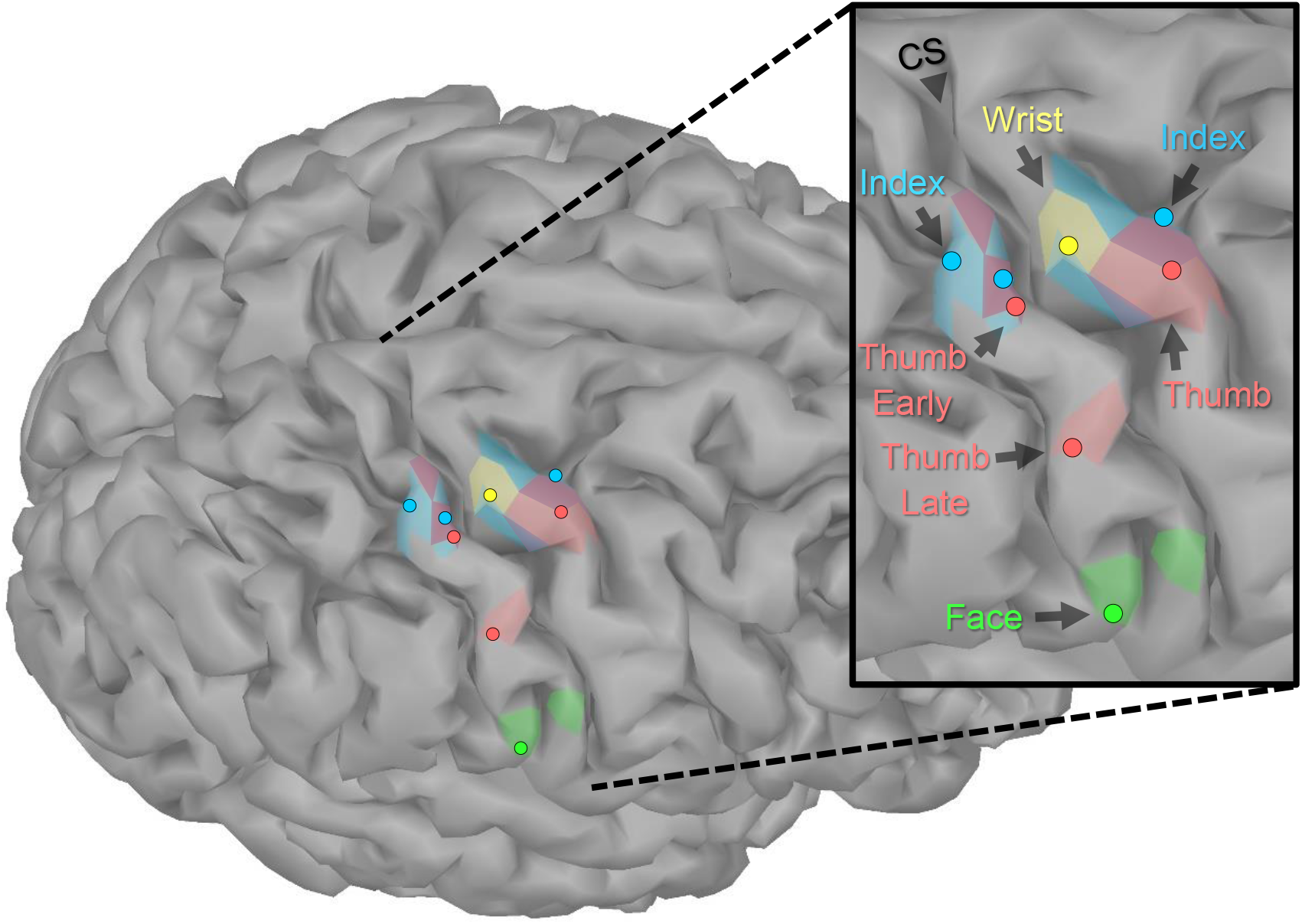
Localization of sensorimotor evoked fields. The peaks of the source activity within the ROIs (shown as dots) showed expected somatotopic organization. The sensory-index task had two equal peaks. The colored regions display the spatial distribution of the source activity within the ROI using manually chosen amplitude thresholds (Sensory: Index 7 pAm, Thumb early 3.5 pAm, Thumb late 5 pAm, Face 20 pAm; Motor: Wrist 8 pAm, Index 6.5 pAm, Thumb 4 pAm). Though the participant had no intact motor or sensory function of his index finger, the expected somatotopic organization was intact. However, index finger maps did overlap with the representations of motor-thumb and wrist and the early component of sensory-thumb.

Figure 1 shows the evoked sensory field waveforms used in the analysis. Somatosensory activity peaks were found near the P35m peak for all body parts. This peak was clear for normal face sensation, but weak for the impaired sensations at the thumb and index finger. However, the source-maps matched what was expected based on anatomy. A late component of the evoked field was observed for the thumb task at 107ms. This activity was also mapped and localized as expected based on anatomy. This later component was not seen on the index finger task, likely because the participant could not perceive the sensation and this late component is related to perception (Hautasaari, Kujala, & Tarkka, 2019; Schubert et al., 2006). However, it should be noted that an earlier component (30 ms) for the index finger had an expected physical location based on anatomy and somatotopy.

**Figure 1:**
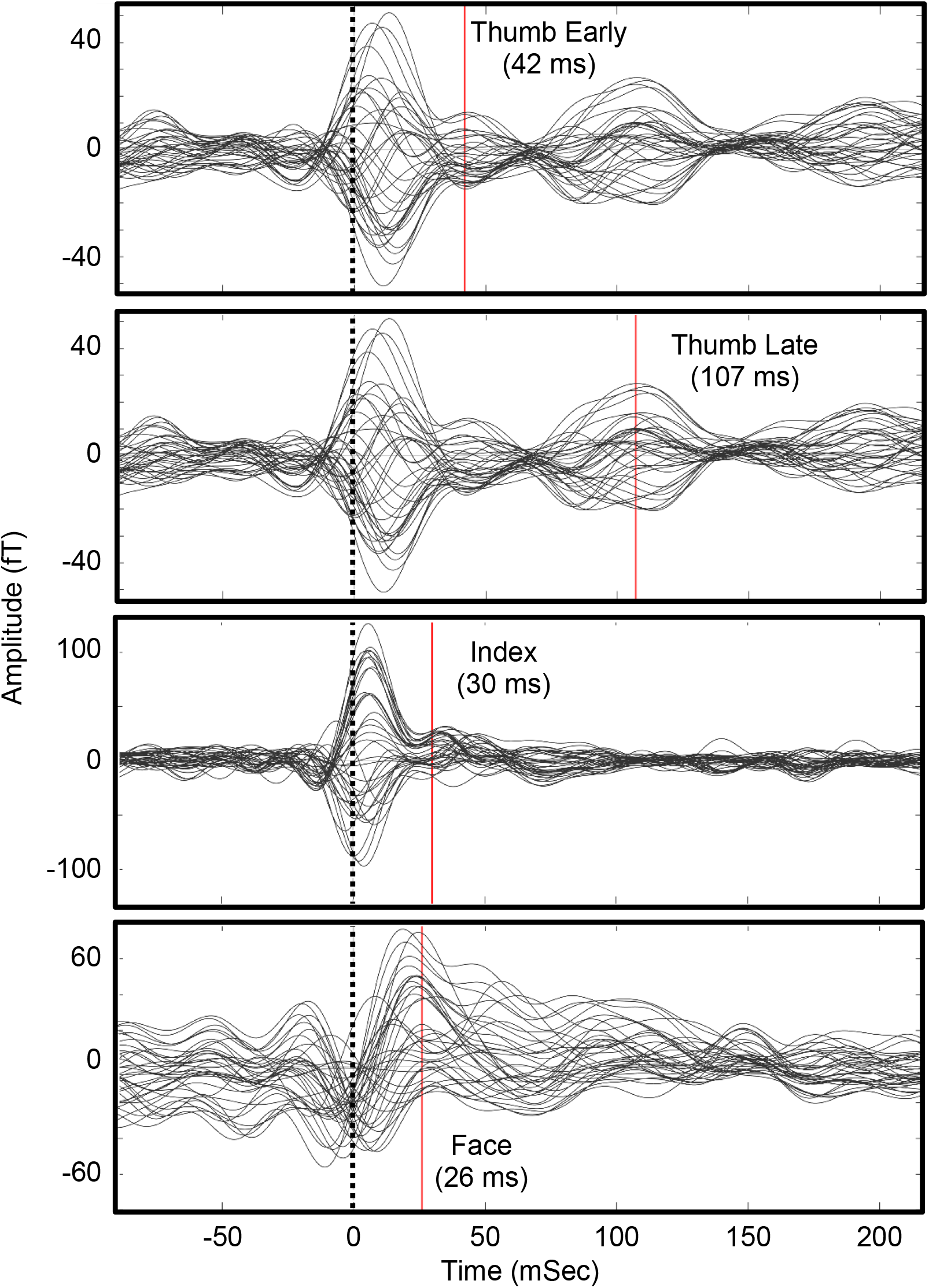
Waveforms for somatosensory evoked fields in the parietal lobe sensors. Times of source analysis are indicated by the red line. An early and late component of thumb stimulation were mapped. An evoked response from the sensory-devoid index finger was difficult to identify.

Figure 2 shows the evoked motor field waveforms used in the analysis. Due to the complexity of the motor tasks, it was challenging to determine the timing for the motor maps. Despite this, the motor activity mapped well to the expected based on somatotopy and anatomy. Typical motor evoke analysis maps the activity before a physical trigger to represent the initiation of the movement. However, this was not possible given the participant’s impairment and the tasks performed. Therefore, the first obvious peak activity in the ROI was mapped. These times were 104 ms after experimenter trigger for thumb (D1) and 67 ms after experimenter trigger for the index finger (D2). These delays fit the expected delays that would occur during a predictable action observation task. As the participant could perform overt wrist extension with limited mobility, the wrist task involved the release of a button to trigger the data by the participant himself. For this wrist task, two components were seen: one just before the trigger (−7 ms) and just after (58 ms). We hypothesize that the initial activity (−7 ms) corresponded to the initiation of the movement where he raised his finger holding down a button while the later component (58 ms) corresponded to the actual execution of the wrist movement. The earlier component mapped to the expected anatomical finger area of the motor cortex (i.e., the inferior part of the hand knob) and overlapped with the index and thumb motor maps. The later component was lateral to the finger areas and the earlier wrist component.

**Figure 2:**
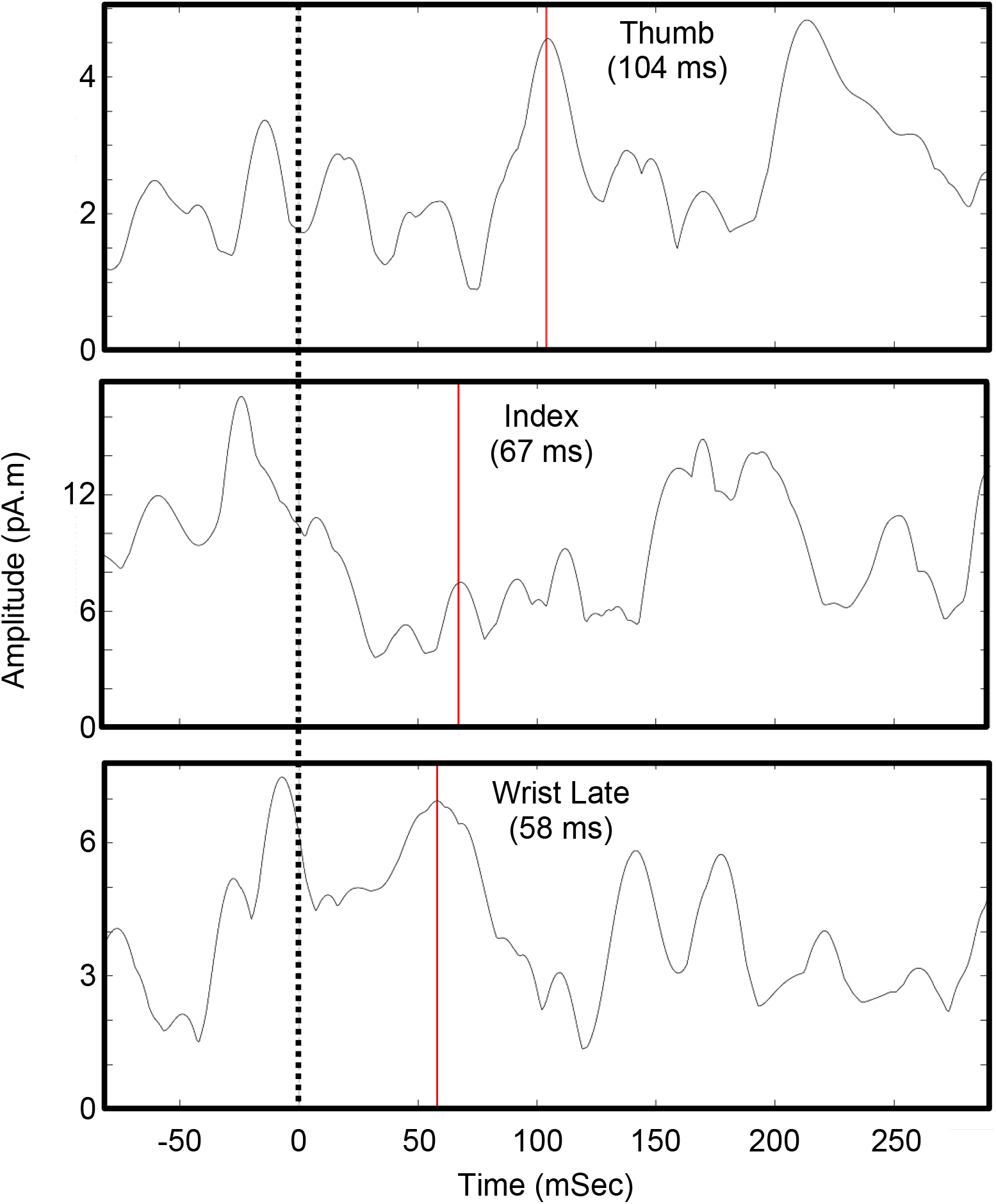
Waveforms for motor evoked fields. Shown is the maximum current density in the pre-central gyrus ROI around the hand knob. Times of source analysis are indicated by the red line. The zero-time point is when a button was pressed by an experimenter (for thumb and index) or released by the participant (wrist).

## 5 Discussion

The evoked fields from impaired somatosensory and motor tasks were successfully mapped for an individual with paralysis and existing implanted BCI hardware. In general, these maps fit the expected anatomy and somatotopic organization (see Figure 3). These maps included both somatosensory and motor function of the index finger despite the participant being unable to elicit any movement or perceive any sensation in that digit. Though the response was modest for the index finger, being able to map body parts without any intact function is encouraging for BCI applications in patients with severe paralysis. Overall, these results add to the limited existing literature that demonstrates that MEG can be used for somatosensory and motor mapping in paralyzed populations. Mapping with MEG is especially critical since mapping with fMRI is often not possible due to implanted metallic objects in this patient population (e.g. fixation, pain stimulators, etc.).

This is the first demonstration of noninvasive sensorimotor mapping in an individual with an existing BCI implant. As BCI technology improves, patients will want to take advantage of new hardware, but it will be important that we can map intact brain activity in those with existing hardware to guide placement of upgrades. It is unlikely that BCI hardware will be MRI compatible in the near future, therefore MEG may play an important role in these cases.

Several key aspects of the methods presented contributed to successful mapping. Having over 700 trials to average across for each impaired task was likely necessary to elucidate the weak evoked fields. To optimize the time with the participant and maximize the number of trials, we had to focus the data collection to a few tasks. To do this, we had to have a deep understanding the participant’s limited intact function and the goal of the mapping (e.g. targeting gyri of the hand area). Another key was noise reduction. Using filtering, such as tSSS, can greatly reduce artifacts, but we still needed to remove 84 sensors due to noise from the previous implant. Also, restricting the analysis to the implant target ROI was necessary since mapping the global peak activity would not have been in the ROI. This is likely due to the artifacts present and/or the weak evoked responses. Mapping more intact functions as intra-subject controls (i.e. wrist movement and face stimulation) confirmed that our data and analysis were appropriate and gave us confidence in the results.

Task design was also important in eliciting the most brain activity associated with impaired function. To map motor function of the completely paralyzed index finger and thumb, our task required the participant to imagine and attempt the movement simultaneously while watching an experimenter performing the movement. Our previous work with participants who were paralyzed due to SCI demonstrated that motor related MEG signals are stronger when attempting to move over just imagining the movement (Foldes, Weber, & Collinger, 2017). Having the participant watch the movement performed also taps into the action-observation and mirror neuron systems (Avanzini et al., 2012; Collinger, Vinjamuri, et al., 2014; Hari et al., 1998; Muthukumaraswamy, Johnson, & McNair, 2004; Neuper et al., 2009; Press et al., 2011).

Interestingly, the two somatosensory peaks for the thumb (42 ms and 107 ms) did not align. The earlier peak was more noisy and right next to the index finger map while the later peak was more pronounced but more lateral. It is difficult to say if this difference is due to noise or if this represents neuroplasticity and remapping/unmasking of thumb representation in the previous index finger representation area. It is possible that the thumb representation expanded medially into the finger area since the participant had only sensation in the proximal part of his thumb. This would suggest that the area between the 42 ms and 107 ms components may be all thumb related. However, the choice to target early or late components for a sensory restoration device implant is not clear. On one hand, the later components may map areas where conscious perception occurs and electrically activating those areas may induce near-normal perceived sensation (Hautasaari, Kujala, & Tarkka, 2019; Schubert et al., 2006). However, stimulating the area of the earlier component may tap into pre-perception areas of the afferent neural stream. This level of detail is speculation. Our implant plan, however, includes three arrays which can cover the two thumb areas and another area of interest such as index finger.

The MEG mapping described here will inform surgical planning and will help minimize the extents of the craniotomy. Awake intraoperative cortical stimulation is also under consideration to further confirm final placement of the electrode arrays. The MEG mapping approach reported here may also be useful in placement of other implant types such as subdural or depth electrocorticography electrodes which have also been demonstrated in evoking focal tactile percepts in the hand (Chandrasekaran et al., 2020; Hiremath et al., 2017).

## Data Availability

Data can be made available upon reasonable request.

